# Uptake of Benue State Health Insurance Scheme among Informal Sector Workers in Makurdi

**DOI:** 10.64898/2026.01.22.26344628

**Authors:** David Tyover Kajo, Chinyere O. Mbachu, Jighjigh Friday Gudu

**Affiliations:** Department of Community Medicine, University of Nigeria, Nsukka

**Keywords:** Health insurance, uptake, informal sector, enrolment

## Abstract

**Introduction:** The provision of affordable and accessible healthcare is a fundamental right that every citizen deserves regardless of their employment and educational status. One way to protect oneself financially from the high cost of receiving medical treatment is through health insurance. This study seeks to determine the perception, willingness to accept/enrol and uptake for Benue State Health Insurance Scheme among informal sector workers in Makurdi.

**Method:** The study was carried out in Makurdi among informal sector workers using a descriptive cross-sectional design. The study population consisted of informal sector workers living in Makurdi. A total of 369 participants were selected using multi-stage sampling technique and data was collected using an interviewer administered questionnaire. The data obtained from the research was examined and analysed using basic percentages and means. Comparative analysis of the study was done by subjecting the data obtained to chi-square analysis using statistical package for social sciences (SPSS), version 23. A P-value less than 0.05 were considered statistically significant.

**Result:** Findings from the study revealed a high willingness to enrol in Benue State Health Insurance Scheme (69.3%) but a low level of enrolment (34.0%) among informal sector workers. The mean willingness to pay amount was found to be 9,020.6 naira per annum as premium. Statistical analysis showed that there was a significant relationship between age, gender, marital status, monthly income, level of education and the willingness to enrol/enrolment in the scheme (p=0.00). The respondents generally had a good perception about BNSHIS as they believed; it will relieve them from making out-of-pocket payment for health care (30.7%), will protect from catastrophic health expenditure (27.9%), will make health care cheaper (39.0%), and will increase access to health services (30.7%). Those unwilling to enrol based their reasons around funds mismanagement (12.6%), alternative means of healthcare (12.6%), lack of regular income (16.2%), lack of trust in the system (24.3%) and non-belief in paying for sickness (13.5%). The less recurrent reasons were lack of interest (9.0%) and do not need health insurance (11.7%).

**Conclusion:** The uptake of Benue State Health Insurance Scheme among informal sector workers in the survey is considerably low. However, most informal sector workers have a good perception about the scheme and are willing to enrol in the scheme. The few who are not willing to enrol based their reasons around funds mismanagement, low income and lack of trust in the system.

## Introduction

The provision of affordable and accessible healthcare is a fundamental right that every citizen deserves regardless of their employment and educational status. Due to high cost of using medical services, worldwide, 1.3 billion people live in underdeveloped nations without access to quality, reasonably priced healthcare^1, 2^. Long wait times, a lack of health facilities, a high cost of out-of-pocket care, and a shortage of medical personnel are further obstacles to accessing health care^3^. While most of developed countries have a prepayment scheme for health, the bulk of individuals who live in underdeveloped nations, particularly in Africa, pay for their own healthcare^4^. When compared to the developed world, this has made the burden of chronic illnesses, disabilities, and death greater and escalated, and has resulted in low productivity, short life expectancies, and poor development^4^.

According to WHO^5^, low-and-middle income countries bear approximately 90% of world’s disease burden, yet only a little percent of global health spending occurs in these countries. Poor administration of public health services, inadequate funding for healthcare and the incapacity of public primary care services to meet the demands of the expanding population are all blamed for this predicament^6^. The challenges often encountered in out-of-pocket expenses have mandated the introduction of prepaid health insurance in many developing countries^7^.

African leaders pledged in 2001 to take all necessary steps to guarantee that resources for healthcare are made available. They decided to provide healthcare improvement 15% of the yearly national budget. The focus of emphasis is now on universal health coverage as a workable way to increase healthcare affordability and accessibility on a global scale. Resolution WHA 58.33, which was passed by the 58th World Health Assembly in May 2005, urged member nations to make sure that their health finance systems contain mechanisms for financial contribution prepayment. These resolutions supported the shift to universal health coverage^8^, believing that social health insurance programs would be an effective tactic for raising health-related funds, sharing risks, giving the underprivileged fair access to healthcare, and producing higher-quality healthcare^3^.

One way to protect oneself financially from the high cost of receiving medical treatment is through health insurance. This is a fundamental component of health care for all. In Nigeria, the implementation of National health insurance has been an essential step towards achieving this UHC.

In 2005, the National Health Insurance Scheme (NHIS) was created with the intention of ‘securing universal coverage and access to adequate and affordable healthcare in order to improve the health status of Nigerians^9^. It pools funds from premium of enrolees and purchased health care through Health Management Organisations (HMOs) registered under the scheme. As of 2021, there were about 60 HMOs registered with the NHIS, 49 (77.6%) of which had nationwide presence^10^. In May 2022 the National Health Insurance Scheme was repealed by the National Health Insurance Authority (NHIA) Act. Despite the creation of the National Health Insurance Scheme (NHIS), most Nigerians are not covered by the scheme^11^. In 2019, it was reported that the scheme covered less than 5% of Nigerians with the enrolees being largely made up of Federal Government employees and their dependents^12^. A survey by the Lagos Bureau of Statistics revealed that only 11% of household members in the State had their healthcare costs covered by any form of health insurance. The NHIS decentralized the health insurance program’s operation in order to close the coverage gap, and state governments were expected to replicate the program in their respective states. But as of 2019, the State-Based Health Insurance Scheme (SHIS) was only enacted by 18 states, with Lagos state being the first to do so in 2015^10^. State governments commit to dedicate a percentage of their consolidated revenue to the scheme to finance premiums for the poor and vulnerable in the state^12^. The decentralization of social health insurance to sub national levels was also an attempt to expand coverage to informal sector, seeing that the approximately 5% that were being covered by the NHIS were formal sector employees.

Insecure employment, poor and irregular income, and self-employment without social protection are the main characteristics of the informal sector. As a result, determining how much money informal sector employees make based on which social security contributions can be subtracted is challenging. Therefore, there are a lot of issues regarding the design of insurance schemes with regard to enrolment, revenue collection, risk pooling, and the purchase of health services that state governments must address if they want to introduce or expand the state social health insurance for the informal sector and to include the poor^13^. This study therefore seek to determine the perception, willingness to accept/enrol and uptake for Benue State Health Insurance Scheme among informal sector workers in Makurdi.

## Methods

### Study Design and area

A descriptive cross-sectional study design was used for this study. The study was carried out among informal sector workers in Makurdi, Benue. Makurdi as the state capital city has an estimated population of 365,000 people and is one of the 23 local government areas of Benue. Makurdi is made up of 11 council wards comprising of both urban and rural dwellings. The major areas which make up its metropolis include; High level, Wurukum, Akpehe, Low level, Wadata, North bank, Kanshio, Gaadi, Agbadu, Fiidi, New GRA, Old GRA. The LGA also has 6 major markets including; Wurukum market, Highlevel market, Wadata market, Modern market, North bank market and international market. Makurdi is home to Benue state university, university of agriculture, Akawe Torkula Polytechnic, Nigeria Army School of Military Engineering, and also, River Benue. There are about 55 health facilities including a federal medical centre and general hospital that are registered with Benue state ministry of health. The people inhabiting the LGA are mostly Tiv, Idoma and Igede, but people of other ethnic groups like Igbo, Hausa and Yoruba also reside in the LGA. Majority are traders, farmers, civil servants, and artisans.

### Study Population

The study population consisted of residents of Makurdi who have been living and working in the study area for at least six (6) months in the informal sector. This population basically includes all males and females of adult age (18years and above).

### Inclusion and exclusion criteria

Adult male and female who resided in Makurdi, work in the informal sector and consented to the study were included in the study.

All males and females below the age of 18, adults who work in the formal sector and those who did not consent to the study were excluded.

### Sample Size Determination

The minimum sample size was determined using Fisher’s formula

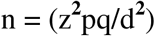

Where n= the desired minimum sample size.

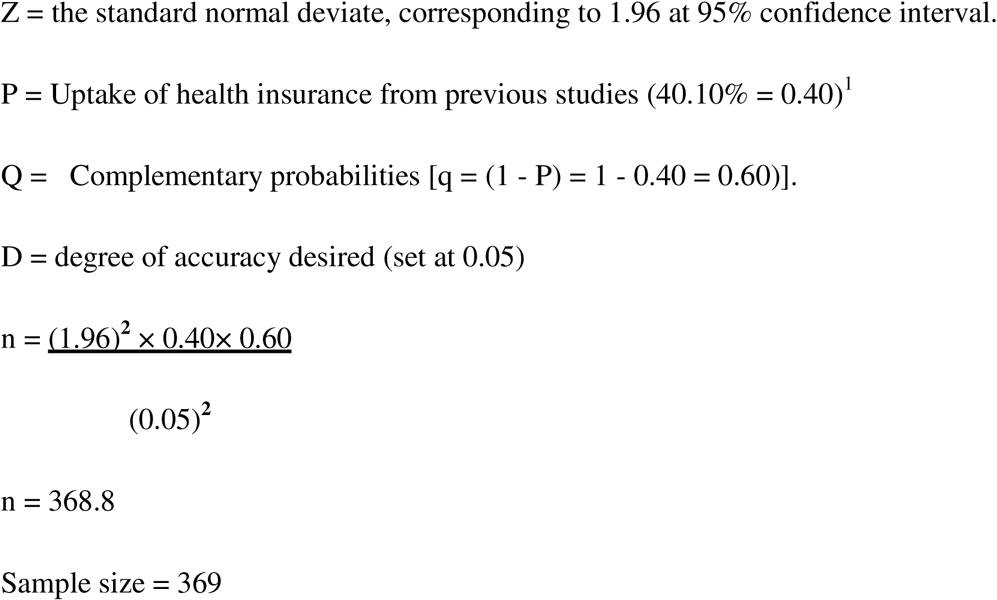

### Sampling Technique

The research sample was chosen using a multi-stage sampling procedure. From the sample frame of 11 wards in Makurdi LGA, 6 wards were chosen by simple random sampling. Simple random sampling (balloting) was used to choose ten streets from each of the wards. Finally, a systematic sampling method was used to select 369 informal workers. In the study, all six major marketplaces were covered.

### Data Collection

Primary data was collected using an interviewer administered questionnaire with open-ended and closed-ended questions. The questionnaire was developed by the researcher after rigorous review of literatures. The questionnaire was pretested and necessary adjustments were made before the final exercise which took a period of two (2) weeks. The questionnaire designed consisted of five sections. Section A contained information on the socio-demographic characteristics of respondents. Section B determined the perception of Benue State Health Insurance Scheme among the respondents. Section C determined willingness to accept (or enrol in the) Benue State Health Insurance Scheme among informal sector workers in Makurdi. Section D determined the level of enrolment of informal sector workers in the Scheme.

### Ethical Consideration

Ethical clearance certificate was sought and obtained from the health research ethics committee of University of Nigeria Teaching Hospital, Enugu and permission to administer questionnaires to patients was obtained from the patients themselves using an informed consent form. The respondents were informed about the objective and purpose of the study, confidentiality was ensured and information was recorded anonymously.

### Data Analysis

Completed questionnaires were collected from respondents and sorted out for ease of computation. The study’s results were examined and analysed using basic percentages and means. Comparative analysis of the study was done by subjecting the data obtained to chi-square analysis using statistical package for social sciences (SPSS), version 23. A P-value less than 0.05 was considered statistically significant.

For Likert scale scoring, means above 3.0 were considered as high acceptability while means below 3.0 were considered as low acceptability. Mean scores were calculated for each variable and cumulative mean was calculated for each domain.

## Results

Out of 369 questionnaires that were administered, 362 were fully completed and analysed, giving a response rate of 98.10%.

Table 1 shows the demographic characteristics of respondents. Results from the study showed that a high number of the respondents were females (51.9% compared to males (48.1%). Higher frequency was observed for married participants (47.5%). This was followed by participants who were single (35.1%), divorced (10.8%) while the least frequency was observed for widowed participants (6.6%). Majority of the participants (53.3%) were between the age ranges of 20-30 years. This was followed by those within the age range of 31-40, above 50 years, below 20 and 41-50 years (25.1%, 16.3%, 16.3% and 11.9% respectively). In terms of ethnicity, Tiv were the majority (40.6%), this was followed by Idoma (26.2%), people from other ethnic groups (16.9%) and traditional Igede (16.3%). Participants with tertiary education recorded higher percentages (36.5%). This was followed by those with secondary education (30.1%), primary education (21.8%) and those with no formal education (11.6%). Majority of the participants were traders (32.9%) while 30.9% were farmers, 22.2% were those with other occupations and 14.1% were artisans.

**Table 1:** Socio-demographic Characteristics of Respondents.

| **Variables** | **Frequency** | **Percentage (%)** |
| --- | --- | --- |
| **Gender** |  |  |
| Male | 174 | 48.1 |
| Female | 188 | 51.9 |
| **Marital Status** |  |  |
| Single | 127 | 35.1 |
| Married | 172 | 47.5 |
| Divorced | 39 | 10.8 |
| Widowed | 24 | 6.6 |
| **Age (years)** |  |  |
| Below 20 | 59 | 16.3 |
| 20-30 | 110 | 30.4 |
| 31-40 | 91 | 25.1 |
| 41-50 | 43 | 11.9 |
| Above 50 years | 59 | 16.3 |
| **Ethnicity** |  |  |
| Tiv | 147 | 40.6 |
| Idoma | 95 | 26.2 |
| Igede | 59 | 16.3 |
| Others | 61 | 16.9 |
| **Level of Education** |  |  |
| No Formal Education | 42 | 11.6 |
| Primary Education | 79 | 21.8 |
| Secondary | 109 | 30.1 |
| Tertiary | 132 | 36.5 |
| **Occupation** |  |  |
| Farmer | 112 | 30.9 |
| Trader | 119 | 32.9 |
| Artisan | 51 | 14.1 |
| Others | 80 | 22.1 |
| **Monthly Income** |  |  |
| <50,000 | 121 | 33.4 |
| 50,000-100,000 | 179 | 49.4 |
| 101,000-150,000 | 45 | 12.4 |
| Above 150,000 | 17 | 4.7 |
| **Total** | **362** | **100.00** |

**Source: Field Survey, (2023)**

In terms of monthly income, majority of the participants were within the income bracket of fifty thousand naira (50,000) to one hundred thousand naira (100,000), this was followed by those with monthly income that is below fifty thousand naira (50,000), one hundred and one thousand naira to one hundred and fifty thousand naira monthly (12.4%) and those who were within the income bracket of above one hundred and fifty thousand naira monthly (4.7%).

The awareness of Benue State health insurance scheme among informal sector workers of Makurdi is presented in table 2. Results obtained revealed that 62.2% of the participants were not aware of Benue State health insurance scheme while 37.8% of the participants were aware of the health insurance scheme. Majority of the participant’s (26.3%) heard of Benue State health insurance scheme from family members and friends. This was followed by those who heard of the scheme on the radio (21.9%), television (19.7%), Newspaper (17.5%), employer (14.6%) and other sources (0.0%).

**Table 2:** Awareness of BNSHIS.

| **Variables** | **Frequency** | **Percentage (%)** |
| --- | --- | --- |
| **Ever heard about Benue State Health Insurance Scheme** |  |  |
| Yes | 137 | 37.8 |
| No | 225 | 62.2 |
| **Do you know how much the premium is** |  |  |
| Yes | 93 | 25.7 |
| No | 269 | 74.3 |
| **Source of your information on Benue SHIS?** |  |  |
| Radio | 30 | 21.9 |
| TV | 27 | 19.7 |
| Newspaper | 24 | 17.5 |
| Employer | 20 | 14.6 |
| Family/Friend | 36 | 26.3 |

**Source: Field Survey, (2023)**

Majority (74.3%) of the participants did not know the insurance schemes’ premium price while 25.7% knew the premium price of the health insurance scheme.

The mean scores for perception of the Scheme among informal sector workers is presented in table 3. The results show that 32.9% and 30.7% of the respondents agreed and strongly agreed that the Scheme will relieve them of OOP for health care, respectively, and the mean score was 3.73 (1.1). With respect to access to health services, 33.4% and 30.7% agreed and strongly agreed, respectively, that the Scheme will enable them to have better access to health services, and the mean score was 3.75 (1.1).

**Table 3:** Perception of Benue State Health Insurance Scheme among Informal Sector Workers in Makurdi.

| **Perception of BNSHIS** | **SD** | **D** | **U** | **A** | **SA** | **$\bar{x}$ (SD)** |
| --- | --- | --- | --- | --- | --- | --- |
| The Scheme will relieve me of out-of-pocket payment | 17(4.7) | 40(11.0) | 75(20.7) | 119(32.9) | 111(30.7) | 3.73(1.1) |
| The Scheme will protect me and my family from high cost. | 27(7.5) | 81(22.4) | 57(15.7) | 96(26.5) | 101(27.9) | 3.45(1.3) |
| The Scheme will provide me a cheaper form of health care | 28(7.7) | 35(9.7) | 80(22.1) | 78(21.5) | 141(39.0) | 3.74(1.2) |
| The Scheme will enable me have better access to health services | 17(4.7) | 37(10.2) | 76(21.0) | 121(33.4) | 111(30.7) | 3.75(1.1) |
| I might be faced with an increased burden of medical bills, if I do not register under the Scheme. | 14(3.9) | 42(11.6) | 53(14.6) | 102(28.2) | 151(41.7) | 3.92(1.1) |
| The Scheme is a waste of money and time | 36(9.9) | 150(41.4) | 94(26.0) | 76(21.0) | 6(1.7) | 2.62(0.9) |
| If I enrol the funds will be mismanaged | 19(5.2) | 88(24.3) | 81(22.4) | 77(21.3) | 97(26.8) | 3.40(1.2) |

**Source: Field Survey, (2023)**

However, considerable proportions of the respondents agreed (21.3%) and strongly agreed (26.8%) that funds will be mismanaged in the Scheme, and the mean score was 3.40 (1.2).

The willingness to enrol and pay for Benue State Health Insurance Scheme is presented in table 4. Results obtained revealed that 69.3% of the participants were willing to accept or enrol in the BNSHIS while 30.7% were not willing enrol in the Benue State health insurance scheme. The mean Willingness to Pay (WTP) amount was found to be 9,020.6 naira. In terms of reasons for unwillingness to enroll in BNSHIS, majority (24.3%) stated that they don’t trust the system will work. This was followed by those stated that lack of regular income (16.2%), don’t believe in paying for sickness (13.5%), afraid of fund mismanagement (12.6%), have other means of healthcare (12.6%), don’t need health insurance (11.7%) and not interested (9.0%) are the reasons for their unwillingness to enroll in the Benue State health insurance scheme.

**Table 4:** Willingness to Enrol and pay for BNSHIS among Informal Sector Workers in Makurdi.

| **Variables** | **Frequency** | **Percentage (%)** |
| --- | --- | --- |
| **Willing to enrol in Benue State Health Insurance Scheme** |  |  |
| Yes | 251 | 69.3 |
| No | 111 | 30.7 |
| **Reason for lack of willingness to enrol in BNSHIS** |  |  |
| I don't trust the system will work | 27 | 24.3 |
| Lack of regular income | 18 | 16.2 |
| I don't believe in paying for sickness | 15 | 13.5 |
| Afraid of funds mismanagement | 14 | 12.6 |
| I have other means of health care | 14 | 12.6 |
| I don't need health insurance | 13 | 11.7 |
| Not interested | 10 | 9.0 |
| **Amount willing to pay annually** |  |  |
| Mean | 9,020.6 |  |
| Standard deviation | 3916.6 |  |

**Source: Field Survey, (2023)**

The level of enrolment of informal sector workers in the BNSHIS scheme is shown in table 5. Results obtained revealed that majority of the participant (66.0%) were not enrolled in the Benue State health insurance scheme, while 34.0% of the participants were enrolled in the scheme.

**Table 5:** Level of Enrollment of Informal Sector Workers in the BNSHIS Scheme.

| Variables | Frequency | Percentage (%) |
| --- | --- | --- |
| **Enrolled in Benue State Health Insurance Scheme** |  |  |
| Yes | 123 | 34.0 |
| No | 239 | 66.0 |
| **Total** | **362** | **100.0** |

**Source: Field Survey, (2023)**

The socio demographic factors influencing willingness to accept or enrol in the Benue State Health Insurance Scheme is presented in table 6. Results obtained revealed that male participants showed the highest willingness (100.0%) to pay/enrol in health insurance, while females showed just 41.0%

**Table 6:** Socio-demographic Factors Influencing Willingness to Enrol in the BNSHIS.

| Variables | Observations | Willingness to Enroll | | $\chi^2$ | P-value |
| --- | --- | --- | --- | --- | --- |
|  |  | Yes (%) | No (%) |  |  |
| **Gender** |  |  |  |  |  |
| Male | 174 | 174 (100) | 0 (0.0) | 148.16 | 0.00 |
| Female | 188 | 77 (41.0) | 111 (59.0) |  |  |
| **Marital Status** |  |  |  |  |  |
| Single | 127 | 127 (100) | 0 (0.0) | 101.03 | 0.00 |
| Married | 172 | 79 (45.9) | 93 (54.1) |  |  |
| Divorced | 39 | 29 (74.4) | 10 (25.6) |  |  |
| Widowed | 24 | 16 (66.7) | 8 (33.3) |  |  |
| **Age (years)** |  |  |  |  |  |
| Below 20 | 59 | 59 (100) | 0 (0.0) | 74.38 | 0.00 |
| 20-30 | 110 | 46 (41.8) | 64 (58.2) |  |  |
| 31-40 | 91 | 75 (82.4) | 16 (17.6) |  |  |
| 41-50 | 43 | 33 (76.7) | 10 (23.3) |  |  |
| Above 50 years | 59 | 38 (64.4) | 21 (35.6) |  |  |
| **Ethnicity** |  |  |  |  |  |
| Tiv | 147 | 147 (100) | 0 (0.0) | 145.20 | 0.00 |
| Idoma | 95 | 26 (27.4) | 69 (72.6) |  |  |
| Igede | 59 | 40 (67.8) | 19 (32.2) |  |  |
| Others | 61 | 38 (62.3) | 23 (37.7) |  |  |
| **Level of Education** |  |  |  |  |  |
| No Formal Education | 42 | 27 (64.3) | 15 (35.7) | 45.49 | 0.00 |
| Primary Education | 79 | 79 (100) | 0 (0.0) |  |  |
| Secondary | 109 | 63 (57.8) | 46 (42.2) |  |  |
| Tertiary | 132 | 82 (62.1) | 50 (35.7) |  |  |
| **Occupation** |  |  |  |  |  |
| Farmer | 112 | 100 (89.3) | 12 (10.7) | 69.76 | 0.00 |
| Trader | 119 | 50 (42.0) | 69 (58.0) |  |  |
| Artisan | 51 | 44 (86.3) | 7 (13.7) |  |  |
| Others | 80 | 57 (71.2) | 23 (28.8) |  |  |
| **Monthly Income** |  |  |  |  |  |
| <50,000 | 121 | 114 (94.2) | 7 (5.8) | 56.51 | 0.00 |
| 50,000-100,000 | 179 | 96 (53.6) | 83 (46.4) |  |  |
| 101,000-150,000 | 45 | 29 (64.4) | 16 (35.6) |  |  |
| Above 150,000 | 17 | 12 (70.6) | 5 (29.4) |  |  |

**Source: Field Survey, (2023)**

All the single participants (100.0%) involved in the study were willing to enroll in the scheme, while their married counterpart showed less willingness (45.9%) to enroll in the scheme. The willingness to enrol/accept the scheme in relation to age revealed that all the participants (100.0%) below 20 years were willing to enrol in the scheme. In terms of ethnicity, all the Tiv participants showed the highest willingness (100.0%) while Idoma showed the least (27.4%). The willingness to enroll/accept the Benue State Health Insurance scheme in relation to level of education revealed that primary school participants were the highest (100%) while secondary were the least (57.8%) willing. For occupation, 89.3% of farmers were willing enroll/accept the health insurance scheme while 42.0% of traders were willing to enroll in the health insurance scheme, showing the least percentage. In terms of monthly income, 94.2% of those whose monthly income is less <50,000 were willing to enroll/accept the program while 53.6% of participants with monthly income bracket of 50,000 – 10,000 were willing to enroll in the scheme. Statistical analysis revealed a significant relationship between gender, marital status, age, ethnicity, level of education, occupation, monthly income and willingness to accept/enrol in the Benue State Health Insurance Scheme.

The socio demographic factors influencing enrolment in the Benue State Health Insurance Scheme is presented in table 7. Results obtained revealed that all the female participants (100.0%) were enrolled in the scheme. Amongst the 174 male participants, 70.7% enrolled while 29.3% of them were not enrolled in the BNSHIS. Single participants showed the highest level of enrolment (73.2%) while widowed showed the least (8.3%). In terms of age, 66.1% of participants who were below 20 years enrolled in the scheme, and were the highest while 7.3% of participants who were within the age range of 20-30years were the least enrolled (7.3%). For ethnicity, Tiv participants showed the highest level of enrolment (74.8%) and Idoma least (0.00%). With regards to level of education, 11.9% of participants with no formal education were enrolled while 88.1% were not enrolled in the health insurance scheme. 72.2% of participants with primary education were enrolled while 27.8% of them were not enrolled in the health insurance scheme.

**Table 7:** Socio-demographic Factors Influencing Enrolment in the BNSHIS.

| Variables | Observations | Enrolled in the Scheme | | $\chi^2$ | P-value |
| --- | --- | --- | --- | --- | --- |
|  |  | Yes (%) | No (%) |  |  |
| **Gender** |  |  |  |  |  |
| Male | 174 | 123 (70.7) | 51 (29.3) | 201.29 | 0.00 |
| Female | 188 | 0 (0.0) | 188(100) |  |  |
| **Marital Status** |  |  |  |  |  |
| Single | 127 | 93 (73.2) | 34 (26.8) | 141.11 | 0.00 |
| Married | 172 | 16 (9.3) | 156 (90.7) |  |  |
| Divorced | 39 | 12 (30.8) | 27 (69.2) |  |  |
| Widowed | 24 | 2 (8.3) | 22 (91.7) |  |  |
| **Age (years)** |  |  |  |  |  |
| Below 20 | 59 | 39 (66.1) | 20 (33.9) | 73.07 | 0.00 |
| 20-30 | 110 | 8 (7.3) | 102 (92.7) |  |  |
| 31-40 | 91 | 45 (49.5) | 46 (50.5) |  |  |
| 41-50 | 43 | 15 (34.9) | 28 (65.1) |  |  |
| Above 50 years | 59 | 16 (27.1) | 43 (72.9) |  |  |
| **Ethnicity** |  |  |  |  |  |
| Tiv | 147 | 110 (74.8) | 37 (25.2) | 187.19 | 0.00 |
| Idoma | 95 | 0 (0.0) | 95 (100) |  |  |
| Igede | 59 | 5 (8.5) | 54 (91.5) |  |  |
| Others | 61 | 8 (13.1) | 53 (86.9) |  |  |
| **Level of Education** |  |  |  |  |  |
| No Formal Education | 42 | 5 (11.9) | 37 (88.1) | 71.75 | 0.00 |
| Primary Education | 79 | 57 (72.2) | 22 (27.8) |  |  |
| Secondary | 109 | 21 (19.3) | 88 (80.7) |  |  |
| Tertiary | 132 | 40 (30.3) | 92 (69.7) |  |  |
| **Occupation** |  |  |  |  |  |
| Farmer | 112 | 50 (44.6) | 62 (55.4) | 56.23 | 0.00 |
| Trader | 119 | 9 (7.6) | 110 (92.4) |  |  |
| Artisan | 51 | 27 (52.9) | 24 (47.1) |  |  |
| Others | 80 | 37 (46.2) | 43 (53.8) |  |  |
| **Monthly Income** |  |  |  |  |  |
| <50,000 | 121 | 79 (65.3) | 42 (34.7) | 84.06 | 0.00 |
| 50,000-100,000 | 179 | 26 (14.5) | 153 (85.5) |  |  |
| 101,000-150,000 | 45 | 14 (31.1) | 31 (68.9) |  |  |
| Above 150,000 | 17 | 4 (23.5) | 13 (76.5) |  |  |

**Source: Field Survey, (2023)**

In terms of monthly income, 65.3% of those whose monthly income is less <50,000 showed the highest level of enrolment. Chi-square analysis revealed that there was a significant relationship between gender, marital status, age, ethnicity, level of education, occupation, monthly income and enrolment in the Benue State Health Insurance Scheme.

## Discussion

This research examined the uptake of Benue State Health Insurance Scheme among informal sector workers in Makurdi. The perception about Benue SHIS is encouraging. Since the individual means are above the decision point of 3.0, it signifies that participants believe that the scheme will ease them of out-of-pocket payment, protect them and their families from high costs, provide them with cheaper form of health care, they can assess primary healthcare, maternal and children health, emergency care and selected specialized services and that it is not a waste of money and time. Other studies^28^ also reported a good perception and a feeling of effectiveness of health insurance schemes on the health and wellbeing of participants, despite challenges. A study by Omotowo et al^25^ also indicates a good perception (59.8%) of health insurance. This positive perception indicates that potential beneficiaries are interested in health insurance; hence a need to scale up the scheme at all levels.

However, participants have a negative perception about the scheme regarding the funds management. They believe that their funds will be mismanaged if they register for Benue state health insurance scheme. The result and findings are comparable to the findings of another study conducted in Enugu metropolis^24^ which showed that 54.1% of the respondents believe the government cannot be trusted to keep its end of the bargain with regards to NHIS.

This study shows that, majority of the respondents were willing to enrol in the Benue SHIS. This is similar to the findings of other studies^29, 25, 2, 60^ which show high prevalence of willingness to enrol or pay for health insurance schemes. According to Ndung’u^3^ and Adewole et al^9^, despite the willingness to participate in health insurance, mainstream policies have not taken the informal sector into account and the majority of those working in the informal sector in low- to middle-income nations lack access to health insurance programs ^3,9^. This perhaps explains the high willingness of the informal sectors workers in Makurdi to enrol in the BSHIS. This finding is a positive indicator that informal sector workers in Makurdi, Benue state are interest towards the state-based insurance schemes, therefore, policy makers and the government are encouraged to scale up resources and improved on the scheme to accommodate more beneficiaries.

Among the participants that were willing to enrol in the BSHIS, over half of them were willing to pay up to 9, 000 for the premium. This corresponds to the findings of the study Akwaowo et al^29^ but lower than the amount reported in the study by Ishaq^59^ which showed that participants were willing to pay as high as 2,626.9 monthly (31,522 naira per annum); as well as with Tabansi et al^2^ where participants were willing to pay as high as much as 70,000 per annum. This however, may be because the participants in Tabansi et al^2^ would rather enrol in a private based scheme.

For the few respondents that were not willing to enrol in the scheme, the reasons they gave are similar to the findings of Osaro et al^31^ and Tabansi et al^2^ which identified lack of interest, non-believe in paying for sickness, had other means of meeting their healthcare needs, poor quality of service, lack of interest, lack of regular income and were afraid of funds mismanagement as their reasons for not willing to pay for SHI. These findings therefore call for more transparency among government and all stake holders in management of funds pertaining to health insurance.

On the level of enrolment, this study shows that majority of the respondents were not enrolled in the Scheme. This corresponds with results of a research conducted in Ghana by Salari et al^61^ which showed 40% coverage rate, and also similar to another study^2^ carried out in Rivers state which showed that 63.0% of the respondents were not enrolled in any health insurance scheme. This low level of enrolment may be due to low level of awareness about the Benue SHIS as findings in the study indicate a low level of awareness among the respondents. Ndung’u^3^ also points out awareness as a determining factor for the level of enrolment in health insurance schemes. The study therefore, recommends that, to encourage potential members to enrol, it is necessary to raise awareness about the presence and health insurance’s worth in comparison to other financial options for healthcare.

In terms of socio-demographic factors influencing, willingness to accept/enrol and enrolment in the Scheme among informal sector workers, the findings of this study shows that socio-demographic factors such as age, gender, marital status, level of income and educational level all influence how informal sector workers in Benue state enrol in the BNSHIS as well as their willingness to enrol. This agrees with the findings of Ngung’u^3^ who conducted research on “factors influencing uptake of national health insurance in the informal sector in Kenya”. Regarding gender, findings from the study further showed that the male participants were more willing (100%) to enrol in BNSHIS than female participants (41%). This however, contradicts with the findings of Ndung’u^3^ which shows female were more willing to enrol in health insurance. Nevertheless, the study agrees with Sabine^40^. A satisfactory percentage of male participants were actually enrolled in the scheme. Higher awareness among male participants may be that male informal sector workers were more informed and aware of the BNSHIS as the source of information sharing about BNSHIS was more common on radio. Males are believed to listen to radio more than females. Because they are so important to community activities relating to immunization of children, reduction of infant mortality, reduction in infectious diseases, and access to hospital deliveries, enrolling females in insurance programs and BNSHIS is crucial. Awareness on the benefits should therefore be improved on to reach more females in the informal sector.

For marital status, participants who were single, widowed or divorced were more willing to enrol in BNSHIS as opposed to the married participants. This is contrary to the findings of Kirigia et al^42^, were the researcher asserted that the necessity to safeguard the children and concern about excessive health spendings may be reasons for married people’s increased demand. This is however, not the case in Benue state. The study therefore recommends more married informal sector workers should be made aware that they can assess primary healthcare, maternal and child health, emergency care and selected specialized services if they enrol in BNSHIS.

In terms of age, the research found that majority of the participants were in their 20s and younger participants were more willing to enrol and also had a greater percentage of enrolment among all the five age groups in the study. The trend was found to be decreasing with age. This is in close agreement with Aboyomi^37^. According to research by Aboyomi^37^, aged farmers in Osun state were less likely to enrol in Nigeria’s National Health Insurance Service (NHIS). The wives of older persons were more likely to have big families and be excluded from the health insurance program. In addition, many elderly people lack the means, training, and drive necessary to enroll in a health plan. The study however, disagrees with the findings of Adebiyi and Adeniji^36^ and that of Ndung’u^3^.

In terms of education, the findings of this study show that participants educational levels had a correlation with their willingness to enrol as well as enrolment in the BNSHIS, which clearly indicates that attaining some level of education has an influence on enrolment and willingness to enrol. This agrees with Mensah and Yeboah^22^. However, participants with primary level of education showed the highest level of willingness to enrol in the Benue SHIS. This is consistent with the findings of Ogundeji et al^32^, who found that respondents with higher educational levels were less likely to be willing to pay for health insurance premiums in their study "to access the factors influencing willingness and ability to pay for SHI in Nigeria." The study offered several explanations for this peculiar propensity, suggesting that people with more education could be better equipped to assess their options. For instance, they could be able to analyze problems with service quality that could have an impact on the overall advantages of signing up for the program. Furthermore, the need for cutting-edge medical technologies and medications for non-communicable diseases is rising, and these needs are frequently not met by benefit packages. As a result, they might have difficulty trusting the system because they are afraid that the benefit bundles will not satisfy their health needs or that they will not get good value for their money^32^

On the level of income, the study discovered that participants with the lowest level of income showed the highest level of willingness to enrol in BNSHIS and also had the highest number of enrolment. The possible explanation to this unusual tendency could be that informal sector workers with lower income may see health insurance as a medium of preventing them from further spending on health once they have subscribed to a premium for one year.

## Conclusion

The uptake of Benue State Health Insurance Scheme among informal sector workers is considerably low. While awareness about BSHIS is also low, most informal sector workers have a favourable perception about the scheme and therefore, showed good attitude towards enrolling in the scheme. The few who are not willing to enrol based their reasons around funds mismanagement, low income, lack of trust in the system. When these factors are successfully dealt with, then the uptake of Benue State Health Insurance Scheme among informal sector workers will greatly improve.

Health insurance is of utmost importance in any community. The burden of out-of-pocket health care costs, especially among most of the population who work in the informal sector, has led to severe health care system inequalities. Based on the findings, this study recommends the need to increase the level of awareness about health insurance among the informal sector. It also recommends that, the state government should consider subsidizing premiums for the impoverished or reassessing yearly premiums to ensure that it is within the financial reach of a larger proportion of workers in the informal sector.

## Data Availability

All data produced in the present study are available upon reasonable request to the authors

## References

1. Agiza, M. M., Adhiambo, O. J., Odongo, O. D., Ogungu, O. D., Awino, O. M., Akoth, O. D. & Atieno, O. H. (2020). Uptake of Health Insurance Schemes Accessible to the Informal Sector Workers in Vihiga Sub county, Vihiga county, Kenya: A cross sectional study: International Journal of Innovative Research and Development. 9(11), 108 – 111. DOI: 10.24940/ijird/2020/v9/i11/NOV20058

2. Tabansi, C. K., Harry, T. & Apugo, U. (2022). Willingness to Enroll in Social Health Insurance and associated factors among Household Heads in Obio/Akpor Local Government Area of Rivers State: Open access, 31(1). 10.1101/2022.12.19.22283656

3. Timothy Theuri Ndung’u (2015). Factrors influencing uptake of national health insurance in the informal sector: A case of Ithanga Division in Murang’A County, Kenya

4. Adewole, D. A., Dairo, M. D. & Bolarinwa, O. A. (2016). Awareness and Coverage of the National Health Insurance Scheme among Formal Sector Workers in Ilorin, Nigeria: African Journal of Biomedical Research, Vol. 19, 2 – 8.

5. WHO (2010). World Health Report:- Health Systems Financing: the path to universal, Geneva.

6. World Bank (2005). Nigeria Health financing system assessment (english).

7. Ibukun, O. A, Olatona, F. A, Oridota, E. S, Okafor, I. P, & Onajole, A. T. (2016). Knowledge and Uptake of Community-Based Health Insurance Scheme among Residents of Olowora, Lagos. Jcs journal, 8 – 10

8. WHO (2005). Sustainable health financing, Universal Health Coverage and Social Health www.who.org on 01/09/2013

9. Adewole, D. A, Akanbi, S. A, Osungbade, K. O, & Bello, S. (2017). Expanding health insurance scheme in the informal sector in Nigeria: awareness as a potential demand-side tool: PanAfrica Medical Journal, 27(52), 2 – 5. doi:10.11604/pamj.2017.27.52.11092

10. Uduu, (2021). Health Insurance in Nigeria – Only 3% of Nigerians are covered. https://www.dataphyte.com/latest-reports/development//health-insurance-in-nigeria-only-3-of-nigerians-are-covered/?amp_markup=1

11. Abiola, A. O., Ladi-Akinyemi, T. W., Oyeleye, O. A., Oyeleke, G. K., Olowoselu, O. I. & Abdulkareem, A. T. (2019). Knowledge and utilisation of National Health Insurance Scheme among adult patients attending a tertiary health facility in Lagos State, South-Western Nigeria: Afr J Prm Health Care Fam Med., 11(1), . 10.4102/phcfm http://www.phcfm.org

12. PricewaterhouseCoopers Limited PWC (2019). Sustainability of State Health Insurance Schemes in Nigeria: Beyong the launch. www.pwc.com/ng

13. Nyorera, E. N & Okibo, W. (2015). Factors affecting uptake of national hospital insurance fund among informal sector workers: a case of nyatike sub-county, kenya. International Journal of Economics, Commerce and Management, UK. 3(3). 2 – 13. http://ijecm.co.uk/

14. Oyefabi, AO., Aliyu, AA. & Idris, A. (2014). Sources of Healthcare Financing among Patients at the Ahmadu Bello University Teaching Hospital, Zaria, Nigeria: Journal of Medicine in the Tropics, 16(1), 27–31.

15. Eboh, A., Akpata, G. O. & Akintoye, A. E. (2016). Health Care Financing in Nigeria: An Assessment of the National Health Insurance Scheme (NHIS): European Journal of Business and Management, 8(27), 24 – 31. ISSN 2222-2839 (Online). www.iiste.org

16. Uzochukwu, B.S.C., Ughasoro, M. D., Etiaba, E., Okwuosa, C., Envuladu, E. & Onwujekwe O. E. (2015). Health care financing in Nigeria: Implications for achieving universal health coverage: Nigerian Journal of Clinical Practice, 18(4), 437 – 443. DOI: 10.4103/1119-3077.154196 www.njcponline.com

17. Odunyemi, A. E. (2021).The Implications of Health Financing for Health Access and Equity in Nigeria. Intech Open. *DOI:* 10.5772/intechopen.98565

18. Canagarajah, S. & Setharaman, S. (2001). Social Protection and the Informal Sector in Developing Countries: Challenges and Opportunities. World Bank Series, World Bank: Washington.

19. Okibo, T. (2015). Influence of health insurance status on childhood cancer treatment outcomes in Kenya. Supportive care in cancer. Springer 28(2).

20. Onasanya, A. A. (2020). Increasing Health Insurance Enrolment in the Informal Economy. JoGH, 10(1), 1 – 3. doi:10.7189/jogh.10.010329 www.jogh.org

21. Ensor, T. & Cooper, S. (2004) . Overcoming barriers to health services: Influencing the Demand Side. Health policy and planning 19(2).

22. Mensah, O. & Yeboah, R. (2022). Assessing the Impact of Education on the Uptake of Health Insurance at the Cape Coast Technical University, Ghana: E-Journal of Humanities, Arts and Social Sciences, 3(12), 556 – 571. 10.38159/ehass.20223124

23. Alesane, A. & Anang, B. T. (2018). Uptake of health insurance by the rural poor in Ghana: determinants and implications for policy: PanAfrican Medical Journal, 31(124): DOI: 10.11604/pamj.2018.31.124.16265 www.panafrican-med-journal.com

24. Okiche, E. L., Okiche, C. Y., Isife, C. T., Obi-Ochiabutor, C. C. & Ogbuabor, C. A. (2021). Health care payment practice, perception and awareness of national health insurance scheme by market women in Enugu Metropolis South East Nigeria. PAMJ, 40(127), https://www.panafrican-med-journal.com//content/article/40/127/full

25. Omotowo, I. B., Ezeoke, U. E., Obi, I. E., Uzochukwu, B.S.C., Agunwa, C. C., Eke, C. B., Idoko, C. A. & Umeobieri, A. K. (2016). Household Perceptions, Willingness to Pay, Benefit Package Preferences, Health System Readiness for National Health Insurance Scheme in Southern Nigeria. Health, 8, 1630–1644. 10.4236/health.2016.814159

26. Olanrewaju, M. F., Ajileye, D. O., Asekun-Olarinmoye, T. F., Adeoye, A. O., Oyerinde, O.O., Adebola, O. & Filade, B. A. (2019). A Study of Percieved Benefit and Barriers towards the Uptake of State Health Insurance Scheme in Lokoja, Kogi State, Nigeria. IOSR Journal of Environmental Science, Toxicology and Food Technology,13(10), 71–76, DOI: 10.9790/2402-1310027176

27. Lawal, N., Safana, M. T., Abdulsamad, H., Bature, M., Maikaka, M. K. & Ihsan, I. S. (2023). Evaluating Enrollee Perceived Benefit of the Contributory Scheme Post Implementation in Katsina State Contributory Scheme North-West of Nigeria. Journal of Quality in Health care & Economics, 6(1), DOI: 10.23880/jqhe-16000318

28. Ajike, S. O., Chinenye-Julius, A. E. & Folarin, M. O. (2020). Evaluating end users’ knowledge and perceived benefit of the National Health Insurance Scheme post implementation in a south-west state of Nigeria. World Journal of Advanced Research and Reviews, 7(3), 85–90, DOI: 10.30574/wjarr.2020.7.3.0338

29. Akwaowo, C. D., Umoh, I., Motilewa, O., Akpan, B., Umoh, E., Frank, E., Nna, E., Okeke, U. & Onwujekwe, O. E. (2021). Willingness to Pay for a Contributory Social Health Insurance Scheme: A Survey of Rural Residents in Akwa Ibom State, Nigeria. Front. Public Health 9:654362. doi: 10.3389/fpubh.2021.654362

30. Bamidele, J. O. & Adebimpe, W. O. (2013). Awareness, Attitude and Willingness of Artisans in Osun State Southwestern Nigeria to Participate in Community Based Health Insurance. Journal of community medicine and primary health, 24(1), 1–11.

31. Osaro, B. O., Jaja, I. D. & Uzosike, T. C. (2021). Awareness and Willingness to Participate in Community Health Insurance Scheme among Household Heads in Rivers State Nigeria. Global Journal of Medical Research, 21(1), Online ISSN: 2249-4618

32. Ogundeji, Y. K., Akomolafe, B., Ohiri, K. & Butawa, N. N. (2019). Factors influencing willingness and ability to pay for social health insurance in Nigeria: PLoS ONE 14(8): e0220558. 10.1371/journal.pone.0220558

33. Onwujekwe, O., Okereke, E., Onoka, C., Uzochukwu, B., Kirigia, J. & Petu, A. (2009). Willingness to pay for community-based health insurance in Nigeria: do economic status and place of residence matter? Health Policy and Planning, 25, 155–16, doi:10.1093/heapol/czp046

34. Ugwu, E. O., Akinboye, D. O., Akinoye, J. I. & Onayemi, O. O. (2013). Predictors of Enrolment in Health Insurance: A Study among Self-employed Workers in Ijebu-ode Local Government Area, Ogun State, Nigeria. Texila International Journal of Public Health, 2520–3134, DOI: 10.21522/TIJPH.2013.08.03.Art001

35. Owoeye, O. G. & Irem, E. J. (2021). A Comparative Assessment of the Knowledge, Enrolment and Factors Affecting the Utilization of National Health Insurance Scheme Among Women Attending Antenatal Care in a Secondary and Tertiary Health Facility in Benin City, Nigeria. Journal of Epidemiological Society of Nigeria, 4(2), 13 – 26.

36. Adebiyi, O. & Adeniji, F. O. (2021). Factors Affecting Utilization of the National Health Insurance Scheme by Federal Civil Servants in Rivers State, Nigeria: The Journal of Health Care Organization, Provision, and Financing, 59(1), 1 – 7. 10.1177/00469580211017626

37. Abayomi, S. O. (2012). Factors influencing households willingness to pay for national health insurance (NHIS) in Osun state, Nigeria. Ethnomed, 6(3), 167–172.

38. Chankova S., C. Atim, & Hatt, L. (2009). Ghana’s National Health Insurance Scheme, in the impact of health insurance in low-and middle –income countries. Brookings institution press. Washington DC.

39. WHO (2008): Closing the gap in a generation .Commission on Social Determinants of Health, Final Report. Retrieved from www.who.org on 01/09/2013

40. Sabine, C. (2012) Gender Equality in Access to Healthcare: The role of social health protection. A case study of RSBY scheme. Giz Discussion Papers.on Social Protection.

41. Bourne, P. A & Moureen Keer-Campbell (2010). Determinants of Self-Rated private Health Insurance Coverage in Jamaica. Health, 2, 541–550

42. Kirigia, J, Sambo, L., Banjamin, N., Germano’Rufaro, C. & Takondwa, M. (2005). Determinants of health insurance ownership among South Africa Women. Bmc health services Research, 5(11).

43. Jangati, Y. (2012). Awareness of health Insurance in Andra Pradesh. International Journal of Scientific and Research Publications, 2(6).

44. Mhere Francis (2013) Health Insurance Determinants in Zimbabwe: A case of Gweru Urban. Journal of Applied Business and Economic, 2(14).

45. Akwasi, K. & Joshua, A. (2013). Effects of Spacial location and Household wealth on health Insurance subscription among women in Ghana: Bmc health service research, 13(221)

46. ILO (2000). Employment and Social Protection in the informal sector .Retreived from www.ilo.org.

47. Sarpong, N., Loag, W., Fobil, J., Meyer, C. G., Adu-Arkodie, Y., May, J. & Schwarz G. (2010). National Health Insurance Coverage and Socio-economic.Status in a rural district of Ghana. Tropical Medicine and International Health, 15(2), 191–197.

48. Adegoriola, A. M. & Omoera, O. S. (2021). Bridging the Information Gap to Keep Educators Healthy: Health Insurance Awareness and Actions by Private School Employees in Nigeria: Research Square, 2 – 10. 10.21203/rs.3.rs-476610/v1

49. Rosenstock, I. M. (1974). Historical Origins of the Health Belief Model. Health Education & Behavior, 2(4), 328–335. doi:10.1177/109019817400200403

50. Janz, N. K. & Becker, M. H. (1984). The Health Belief Model: A Decade Later. Health, 11(1): 1–47.doi: 10.1177/109019818401100101

51. Dzulkipli, M. R., Maon, S. N. & Kamarudin, N. H. (2019).Predicting Intention to Purchase Medical and Health Insurance using The Health Belief Model: International Tourism and Hospitality Journal, 2(3), 1 – 7. Online ISSN: 2616-4701 https://rpajournals.com/ithj

52. Lattof, S. R. (2018). Health Insurance and Careseeking Behaviours of Female Migrants in Accra, Ghana. Health Policy Plan, 0(0): 1–11. 10.1093/heapol/-czy012

53. Prakoso, A. D., Sulaeman, E. S. & Suryono, A. (2020). Application of Health Belief Model on Factors Affecting Participation in the National Health Insurance Scheme among Informal Sector Workers in Kudus, Central Java: Journal of Health Policy and Management, 5(1), 61–72. 10.26911/thejhpm.2020.05.01.06 www.thejhpm.com

54. Brahmana, R., Brahmana, R. K. & Memarista, G. (2018). Planned behaviour In purchasing health Insurance: The South East Asian Journal of Management. DOI:10.21002/seam.v12i1.7465

55. Porter, M. E. (2010). What is value in health care? The New England Journal of Medicine, 363, 2477–2481

56. Zelizer, V. A. (1978). Human values and the market: The case of life insurance and death in. American Journal of Sociology, 84(3), 591–610.

57. Begg, D. S., Fischer, R. & Dornbusch (2002). Economics. Health Financing: Designing and Implementing Pro-Poor Policies (London: London: The McGraw-Hill, DFID Health Systems Resource Centre).

58. Cameron, A. C., Trivedi, P., Milne, F. & Piggott, J. (1988) A Microeconometric Model of the Demand for Health Care and Health Insurance in Australia. Review of Economic Studies LV.

59. Ishaq, K.Y. (2022). Willingness to pay for Social Health Insurance Among Informal and Private Sector Workers in Kano Metropolis: A Contingent Valuation Study. International Journal of Social Sciences and Humanities Reviews, 12(1), 495 – 501.

60. Cheno, R.W., Tchabo, W. & Tchamy, J. (2021). Willingness to join and pay for community-based health insurance and associated determinants among urban households of Cameroon: case of Douala and Yaounde. Science direct. 10.1016/j.heliyon.2021.e06507.

61. Salari, P., Akweongo, P., Aikins, M. & Tediosi, F. (2019). Determinants of health insurance enrolment in Ghana: evidence from three national household surveys. Health Policy and Planning, 34, 582 – 594. doi: 10.1093/heapol/czz079

